# Evidence for SARS-CoV-2 Spike Protein in the Urine of COVID-19 patients

**DOI:** 10.1101/2021.01.27.21250637

**Authors:** Santosh George, Anasuya Chattopadhyay Pal, Jacqueline Gagnon, Sushma Timalsina, Pallavi Singh, Pratap Vydyam, Muhammad Munshi, Joy E. Chiu, Isaline Renard, Christina A. Harden, Isabel M. Ott, Anne E. Watkins, Chantal B.F. Vogels, Peiwen Lu, Maria Tokuyama, Arvind Venkataraman, Arnau Casanovas-Massana, Anne L. Wyllie, Veena Rao, Melissa Campbell, Shelli F. Farhadian, Nathan D. Grubaugh, Charles S. Dela Cruz, Albert I. Ko, ‘Yale IMPACT study’ team, Amalia Berna Perez, Elikplim H. Akaho, Dennis G Moledina, Jeffrey Testani, Audrey R John, Michel Ledizet, Choukri Ben Mamoun

## Abstract

SARS-CoV-2 infection has so far affected over 42 million people worldwide, causing over 1.1 million deaths. With the large majority of SARS-CoV-2 infected individuals being asymptomatic, major concerns have been raised about possible long-term consequences of the infection. We developed an antigen capture assay to detect SARS-CoV-2 spike protein in urine samples from COVID-19 patients whose diagnosis was confirmed by PCR from nasopharyngeal swabs (NP-PCR+). The study used a collection of 233 urine samples from 132 participants from Yale New Haven Hospital and the Children’s Hospital of Philadelphia obtained during the pandemic (106 NP-PCR+ and 26 NP-PCR-) as well as a collection of 20 urine samples from 20 individuals collected before the pandemic. Our analysis identified 23 out of 91 (25%) NP-PCR+ adult participants with SARS-CoV-2 spike S1 protein in urine (Ur-S+). Interestingly, although all NP-PCR+ children were Ur-S-, 1 NP-PCR-child was found to be positive for spike protein in urine. Of the 23 Ur-S+ adults, only 1 individual showed detectable viral RNA in urine. Our analysis further showed that 24% and 21% of NP-PCR+ adults have high levels of albumin and cystatin C in urine, respectively. Among individuals with albuminuria (>0.3 mg/mg of creatinine) statistical correlation could be found between albumin and spike protein in urine. Together, our data showe that 1 of 4 of SARS-CoV-2 infected individuals develop renal abnormalities such as albuminuria. Awareness about the long-term impact of these findings is warranted.

## INTRODUCTION

December 31, 2019 will be remembered in the history of humanity and the annals of medicine, epidemiology and global health as the D-day of the ongoing coronavirus pandemic. It is the day when health authorities in Wuhan city, China reported a cluster of patients with symptoms of unidentified pneumonia, with characteristics reminiscent of viral pneumonia.^1-4^ By March 12, 2020, the disease spread across the globe with 145,200 cases, leading the World Health Organization to declare COVID-19 a global pandemic.^5^ As of mid-January 2021, more than 96 million cases and over 2 million deaths have been recorded worldwide. Epidemiological estimates suggest that approximately 70% of the world’s population could become infected with COVID-19, with global case fatality rate between 0.1% and over 25%.^6^

Coronavirus-related illnesses in humans are caused by seven viruses, four of which (HCoV-229E, HCoV-OC43, HCoV-NL63 and HCoV-HKU1) cause common cold symptoms and three others (SARS-CoV, SARS-CoV-2 and MERS-CoV), which cause severe acute respiratory syndrome (SARS) and Middle East respiratory syndrome (MERS), respectively. SARS-CoV and MERS-CoV are known to have fatal outcomes, as occurred during the two outbreaks reported in 2002/2003 and 2012, respectively.^7, 8^ SARS-CoV-2, the agent of COVID-19 pandemic, is a member of betacoranaviruses, which mostly infect bats.^9^ However, this enveloped, positive-stranded RNA virus is also capable of infecting humans.^10^ Genetic characterization of SARS-CoV-2 shows it is closely related to bat virus RaTG13.^11^

Similar to the extensively studied SARS-CoV,^12, 13^ the spike (S) protein of the SARS-CoV-2 plays a crucial role in viral attachment to the human cell surface membrane angiotensin-converting enzyme 2 (ACE2) receptor and entry into the target cell.^14, 15^ This process is accompanied by proteolytically activating the SARS-CoV-2 spike protein at the S1/S2 site, by cleaving S1 from S2 protein by host proteases.^15^ Therefore, screening for SARS-CoV-2 spike protein could be an excellent strategy to monitor active and recent COVID-19 infections. Analysis of the tissue distribution of ACE2 showed high expression of the receptor in the epithelial cells of intestine, renal, alveolar, heart, artery, and gastrointestinal system,^16^ suggesting that in addition to the lungs and the upper respiratory tract, SARS-CoV-2 could also invade other important organs including the kidneys and cause inflammation with possibly long lasting injuries.^17^ To date, the impact of SARS-CoV-2 infection on renal function has been controversial. Analysis of a cohort of 193 adult patients with laboratory confirmed SARS-CoV-2 infection from 3 hospitals in and around Wuhan, China showed kidney abnormalities (CT scan) with 63% of the participants showing proteinuria.^18^ In the USA, a study by Hirsch *et al*., showed that 37% of COVID-19 patients developed Acute Kidney Infection (AKI) during hospitalization, most of whom were on mechanical ventilation.^19^ However, another study by Wang *et al*., on 116 COVID-19 confirmed patients from Renmin Hospital in Wuhan, China found that AKI was uncommon, and that SARS-CoV-2 infection did not result in AKI or exacerbate Chronic Kidney Disease.^20^ Although these studies focused on the presence of specific biomarkers of kidney injury, analysis of urine for the presence of SARS-CoV-2 proteins in COVID-19 patients or asymptomatic individuals have not been examined heretofore.

In this study, we developed an antigen capture assay that detects the presence of SARS-CoV-2 spike protein in biological specimens and used it to evaluate the presence of the antigen in urine samples from nasopharyngeal (NP) PCR positive (NP-PCR+) and negative (NP-PCR-) adults and children collected during the COVID-19 pandemic. Our study population also included urine samples collected 3 to 5 years prior to the pandemic. We found that in our study population, 25.2% of adult COVID-19 patients had SARS-CoV-2 spike S1 protein in their urine at least once during the course of infection. Urine protein analyses further revealed proteinuria with elevated albumin and cystatin C in 24% and 21% of NP-PCR+ individuals, respectively.

## MATERIALS AND METHODS

### Definitions and Calculations

A urine sample was considered positive for SARS-CoV-2 spike protein if the optical density (OD) value obtained was greater than the OD value obtained by COVID-19 NP-PCR-urine sample spiked with 5ng/mL SARS-CoV-2 spike protein. Levels of SARS-CoV-2 spike protein, albumin and cystatin C in urine were normalized to urine creatinine levels and expressed as mg/mg of urine creatinine.^21, 22^ The fractional excretion of sodium (FENa) was calculated using the formula^23-25^

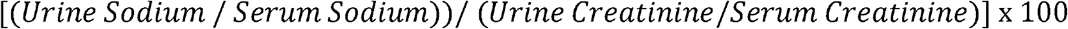

The FEUrea was calculated using the formula ^23-25^

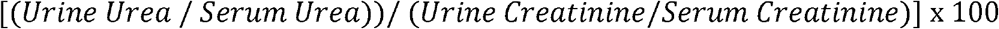

### Study Population

A total of 253 urine samples from 152 participants were analyzed for the purpose of this study. Out of these, 233 urine samples were collected between March and August 2020 (COVID-19 period; N=132) and 20 urine samples were collected before December 2019 (Pre-COVID-19 period; N=20). Participants from the COVID-19 period were recruited from both Yale-New Haven Hospital (YNHH; COVID-19 NP-PCR positive Inpatients, N=91; COVID-19 NP-PCR negative healthcare workers, N=13) and Children’s Hospital of Philadelphia (CHOP; COVID-19 NP-PCR positive children, N=12; COVID-19 NP-PCR negative healthy children, N=14). In addition, two adult NP-PCR-participants were included in the study. A participant was considered positive for COVID-19 by performing SARS-CoV-2 RT-qPCR on nasopharyngeal swabs (NP-PCR) as previously described.^26^

Samples from the Pre-COVID-19 period used as controls included Yale Kidney BioBank participants (N=10), Heart Failure (HF; N=5) and healthy participants (N=5) were served as controls. These samples were collected between 2015 – 2018. Convenience sampling technique was adopted where no statistical methods were used to predetermine sample size. A flowchart describing the study population is shown in Figure 1A.

**Figure 1.**
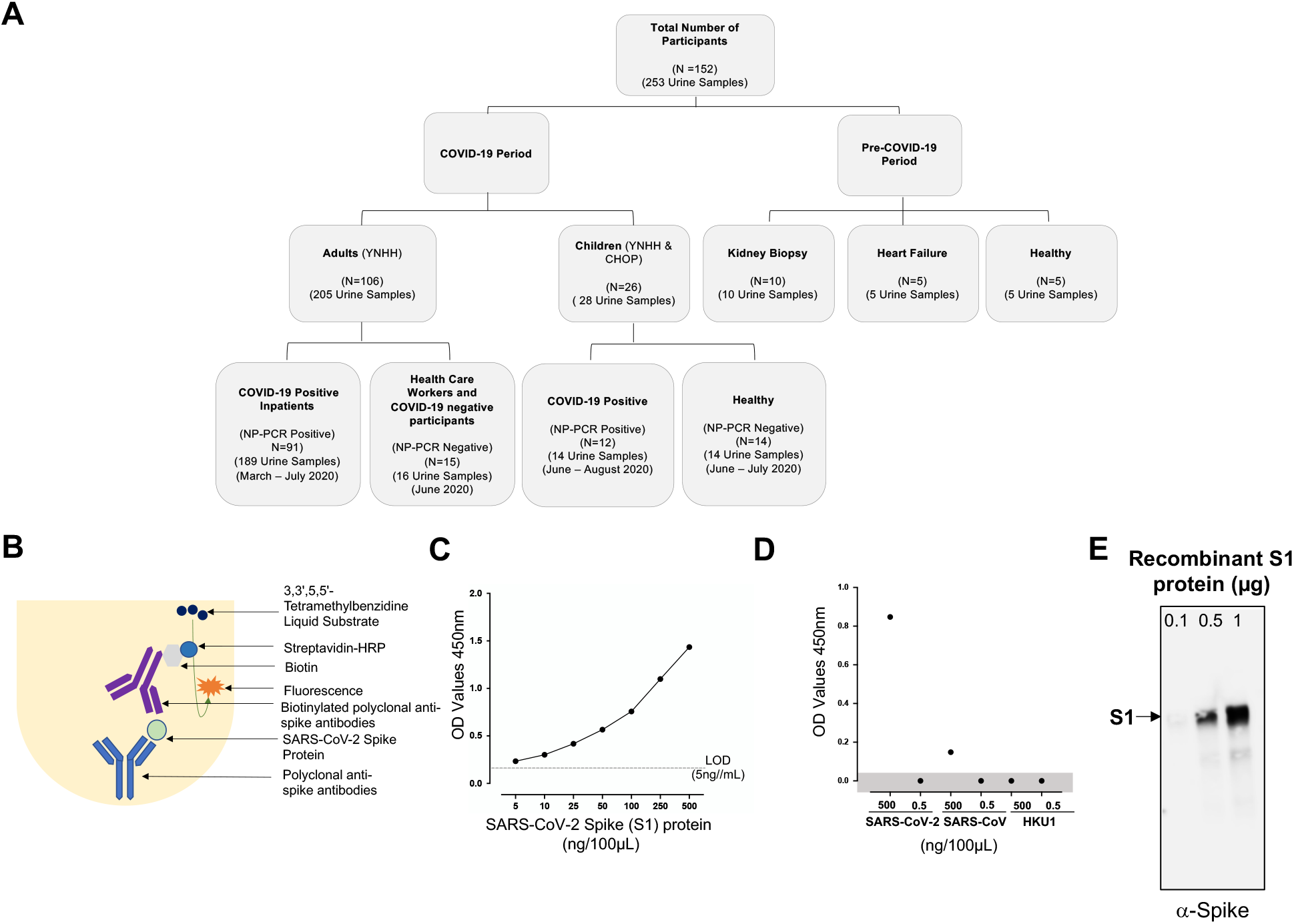
Consolidated summary of study population, assay chemistry, and sensitivity and specificity of the SARS-CoV-2 spike protein using capture ELISA. (A) Flowchart describing the study population. Samples used in this study were collected both before and during the COVID-19 pandemic. (B) Schematic representative of the Capture ELISA assay chemistry. (C) Representative standard curve generated using 5µg/mL SARS-CoV-2 polyclonal anti-spike antibodies. (D) Assay to define the specificity of the SARS-CoV-2 Capture ELISA. Two different concentrations (5µg/mL and 5ng/mL) of different coronavirus infecting humans (SARS-CoV-2, SARS-CoV and HCoV-HKU1) were assessed to determine the specificity of the Polyclonal anti-spike SARS-CoV-2 antibodies. Data points in the shaded area are below the limit of detection. (E) Sensitivity of the polyclonal antibodies to detect SARS-CoV-2 spike S1 protein using Western blot. SARS-CoV-2 in three different concentration was measured 0.1µg, 0.5µg and 1µg.

### Sample processing, RNA extraction and qPCR detection

Urine samples were centrifuged at 2000 RCF for 10 min at room temperature and immediately used for RNA extraction or stored frozen at −80°C. RNA extraction and RT-qPCR detection for SARS-CoV-2 in urine followed the procedures detailed in reference.^26^ From CHOP, all urine samples were treated with NP40 for viral inactivation prior to shipping to Yale School of Medicine. Our comparative analysis using Capture ELISA on urine samples from NP40 treated and non-treated samples, revealed no-effect of NP40 on the assay sensitivity.

### Detection of SARS-CoV-2 spike protein using Urine Capture ELISA (U^ELISA^) and Serum Capture ELISA (S^ELISA^)

Rabbit polyclonal anti-spike protein antibody purified using Protein G immunoaffinity chromatography was purchased from MyBioSource (MBS434243; 2µg/µL) and used as capture antibody in the capture ELISA. This antibody was also biotinylated using EZ-Link™ Micro Sulfo-NHS-Biotinylation Kit (ThermoFisher; Cat No 21925) and used as detection antibody. In addition, SARS-CoV-2 spike protein S1 (78.3 kDa) obtained from GenScript (Cat No Z03501) was used as a positive control (Figure 1B). For the urine capture ELISA (U^ELISA^), urine samples from participants who were NP-PCR-were spiked with different concentrations of SARS-CoV-2 S1 protein (500ng/100µL; 250ng/100µL; 100ng/100µL; 50ng/100µL; 25ng/100µL; 10ng/100µL; 5ng/100µL; 0.5ng/100µL and 0.05ng/100µL; Figure 1C) and used as a standard in every plate. For the serum capture ELISA (S^ELISA^), human serum sample from a healthy individual was spiked with different concentrations of SARS-CoV-2 S1 protein (500ng/100µL; 250ng/100µL; 100ng/100µL; 50ng/100µL; 25ng/100µL; 10ng/100µL; 5ng/100µL; and 0.5ng/100µL; Figure S4). A 96 ELISA well-plate (Nunc MaxiSorp Plate; ThermoFisher Cat No 442404) was coated with 5µg/mL polyclonal anti-spike antibody diluted in carbonate coating buffer (0.848g Na_2_CO_3_, 1.428g NaHCO_3,_ 500mL distilled water) and incubated at room temperature for 2 hours. Unbound antibodies were removed, and 300µL of blocking solution (PBS 0.05% Tween-20 (PBST)-2%BSA) was added per well. The plate was then incubated for an hour at room temperature. In case of U^ELISA^, this was followed by an addition of 100µL of urine sample per well to screen for the presence of SARS-CoV-2 spike protein and the plate was incubated over night at 4°C. In case of S^ELISA^, this was followed by the addition of 20µL of serum sample and 80µL of PBS per well to screen for the presence of SARS-CoV-2 spike protein and the plate was incubated over night at 4°C. After 15-16 hours, the wells were washed four times with PBST, biotinylated polyclonal anti-COVID-19 antibodies (spike protein) were added at a concentration of 5µg/mL, and plates were incubated for 1 hour at room temperature. After four washes with PBST, HRP-streptavidin conjugate (Seracare Life Sciences INC; Cat No KPL 474-3000) was added at a dilution of 1:10000 in PBST, and plates were incubated for 1 hour at room temperature. A final wash (4X) with PBST was then carried out, before adding 100µl of 3,3’,5,5’-Tetramethylbenzidine Liquid Substrate (SureBlue Reserve TMB 1-Component Microwell Peroxidase Substrate, Seracare Life Sciences INC; Cat No KPL 5120-0083) to each well. The optical density (OD) was measured with BioTek FLx800 fluorescence plate reader at 450nm. A schematic of the antigen capture ELISA is shown in Figure 1B.

In addition, to determine the specificity of polyclonal SARS-CoV-2 anti-spike antibodies (MyBioSource; MBS434243; 2µg/µL), different concentrations (0.1µg, 0.5µg and 1µg) of purified recombinant SARS-CoV-2 S1 (GenScript; Cat No Z03501) were probed using a Western blot (Figure 1E).

### Evaluation of the specificity of the detection of SARS-CoV-2 spike protein by urine capture ELISA

Two recombinant proteins, SARS-CoV spike protein (BEI Resources, NR-722, Lot No 660P029) and HCoV-HKU1 spike glycoprotein (BEI Resources, NR-53713, Lot No 70037425) were obtained from BEI Resources, NIAID, NIH. These proteins were used to spike NP-PCR-urine sample at 5µg/mL and 5ng/mL in the urine capture ELISA. In addition, two NP-PCR+ urine samples were spiked with SARS-CoV-2 Spike (S1) protein at the same concentrations as stated above. The cross-reactivity of polyclonal anti-spike antibody was measured in optical density (OD) at 450nm.

### Urine electrolyte, albumin and cystatin C analyses

Urine electrolytes were measured using ion sensitive electrodes on the Randox Imola clinical chemistry analyzer (Randox Laboratories, Ireland, UK). Urine creatinine was determined using a modified Jaffe method. Creatinine measurements are standardized to National Institute of Standards and Technology reference material (SRM 967). Cystatin C and microalbumin were used in accordance with the manufacturer’s instructions (Randox Laboratories, Crumlin, UK).

### Determining the integrity of SARS-CoV-2 spike S1 protein using SDS-PAGE

To determine the integrity of SARS-CoV-2 spike protein in urine, 10µL urine samples from NP-PCR+, NP-PCR- and KB participants were analyzed on a 4-20% MINI-PROTEAN TGX gel (Bio-Rad, USA Cat No 4568096). The gel was analyzed by Western blot after transfer to nitrocellulose membrane (Bio-Rad, USA, Cat No 1620214). The membrane was blocked with 5% milk (American Bio Cat No AB10109-00100) in PBST followed by treatment with polyclonal antibody against SARS-CoV-2 spike (S1+S2) protein raised in rabbit (MyBioSource MBS434243) and used at 1:1000 dilution in PBST. Goat anti-rabbit IgG horse radish peroxidase (HRP) conjugate (Thermofisher Scientific, USA, Cat No 31466) was used as the secondary antibody at 1:5000 dilution. Following this, the membrane was treated with SuperSignal West Pico PLUS chemiluminescent substrate (Thermo Scientific, USA, Cat No 34577) and scanned and imaged using Odyssey fc (Li-cor Biosciences; Cat No 2800-03).

### Statistical analyses

Continuous variables are expressed as either mean [95%CI] or median [IQR]. Statistical analyses were performed using a two-tailed unpaired Student’s t-test or one-way Analysis of Variance (ANOVA) in case of multiple variables. Categorical variables are expressed as numbers (%). Differences were considered statistically significant when p < 0.05.

## RESULTS

### Identification of SARS-CoV-2 spike protein in urine and demographic characteristics

Large scale screening of SARS-CoV-2 positive individuals to identify both symptomatic and asymptomatic individuals is a major priority in the control of COVID-19 disease transmission worldwide. This screening may be facilitated by the use of easy-to-collect biospecimens over several days during the course of an infection or following suspected exposure. To detect the presence of SARS-CoV-2 spike protein in biological specimens from Yale-New Haven Hospital (YNHH) and Children’s Hospital of Philadelphia (CHOP) (Figure 1A, Table 1), we developed a capture ELISA using anti-spike polyclonal antibodies for antigen capture and biotinylated antibodies for detection of the antigen-antibody complex (Figure 1B). In order to use this sandwich ELISA for detection of SARS-CoV-2 spike protein in a collection of urine samples, the assay was optimized using purified recombinant S1 antigen in PBS (data not shown) and urine samples from confirmed NP-PCR SARS-CoV-2 negative individuals and standard curves were generated (Figure 1C).

**Table 1.**
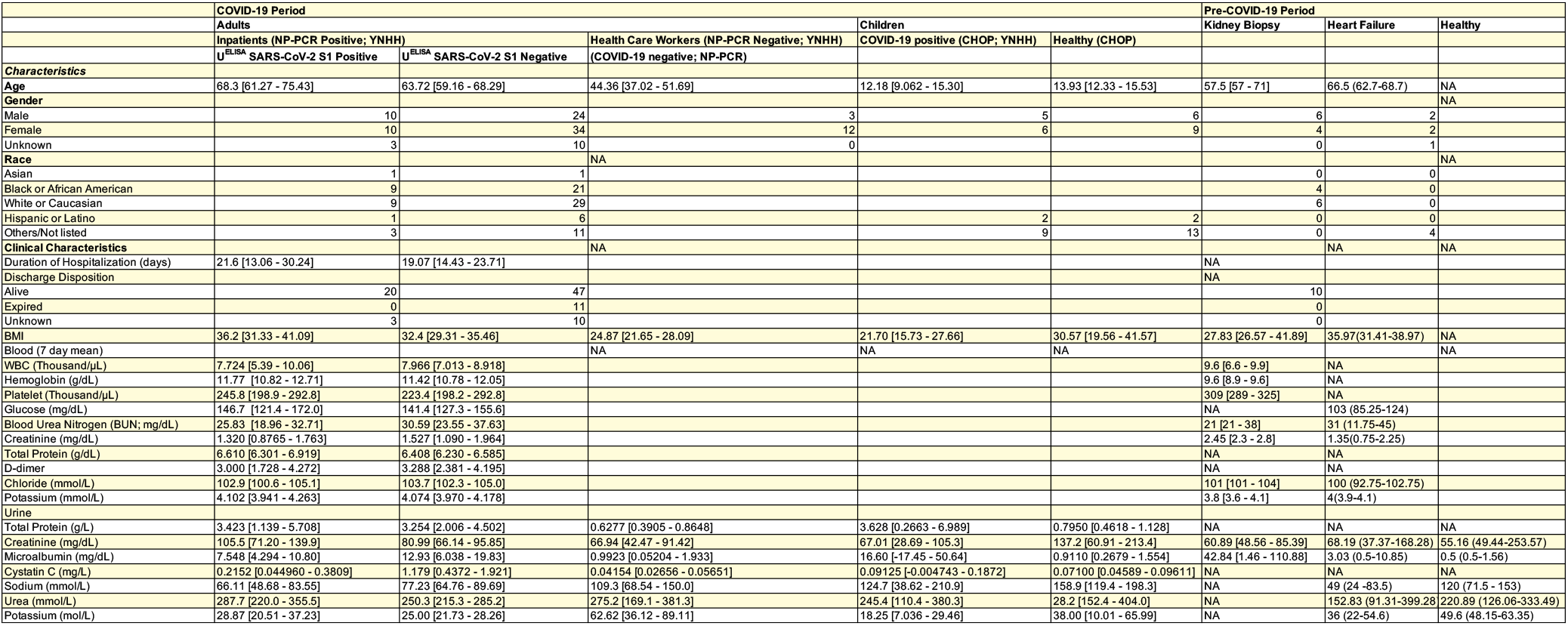
Clinical and demographic characteristics of the study participants.

Using this assay, we determined the lower limit of detection (LOD) to be 5 ng/mL of urine (Figure 1C). The specificity of the antibodies was further evaluated by ELISA using available recombinant spike proteins from SARS-CoV and HCoV-HKU1. While no cross-reactivity could be detected with HCoV-HKU1 spike protein at concentrations as high as 5 µg/mL or with SARS-CoV spike protein at 5 ng/mL, a weak signal could be detected by ELISA with SARS-CoV spike protein at 5 µg/mL (Figure 1D). Western blot analysis confirmed the specificity of the antibodies towards SARS-CoV-2 spike protein (Figure 1E).

Having determined the sensitivity and specificity, the assay was then used to examine a repository of 253 urine samples collected from 152 participants for the presence of SARS-CoV-2 spike protein (Figure 1A; Table 1). This repository included 203 urine samples from SARS-CoV-2 NP-PCR+ adults (189 urine samples; N=91) and children (14 urine samples; N=12) from YNHH and CHOP. Samples from COVID-19 NP-PCR-healthcare workers (HCW; N=13) and children (N=12) from both hospitals, adult COVID-19 negative participants (N=2), as well as urine samples collected prior to the COVID-19 pandemic (N=20) were used as controls (Figure 1A). Creatinine levels in all the urine samples were measured and used to calculate the urine protein-to-creatinine ratio in order to standardize measurements. Of the 203 urine samples from 103 COVID-19 NP-PCR+ adults and children analyzed in this study, 29 (N=23; 25.2%) were found to have SARS-CoV-2 spike protein in the urine (Figure 2A). Interestingly, one child (with no respiratory symptoms) who was negative for SARS-CoV-2 by NP-PCR (performed for pre-admission screening) appeared to have high levels of SARS-CoV-2 spike protein in urine (Figure 2A). Overall, the concentration of SARS-CoV-2 spike protein in adults was 0.033 [0.0096 – 0.0564] (mean [95% CI]; mg/mg of urine creatinine), while the child had 0.0083 (mg/mg of urine creatinine). None of the urine samples from adult HCW (N=15; YNHH), NP-PCR+ children (N=12) or Pre-COVID-19 participants (N=20) showed the presence of SARS-CoV-2 spike protein (Figure 2A). No correlation between the presence of SARS-CoV-2 spike protein in the urine and the gender of the COVID-19 patient could be found (P=0.34; Figure 2B) among 20 of the 23 participants with SARS-CoV-2 spike protein in their urine (information on gender for three individuals was not available). Similarly, no significant association between the presence of the SARS-CoV-2 spike protein and factors such as body mass index (BMI; P=0.16), age (P=0.29) and duration of hospitalization (P=0.49) could be found (Figure 2C). In this cohort, we also examined possible correlations between the levels of albumin and cystatin C in the urine samples and BMI, age and duration of hospitalization of the patient. However, no significant association was observed between these confounding factors and elevated levels of albumin (BMI P=0.33, age P=0.058, duration of hospitalization P=0.12) and cystatin C (BMI P=0.36, age P=0.88, duration of hospitalization P=0.11) (Figure S3A and S3B).

**Figure 2.**
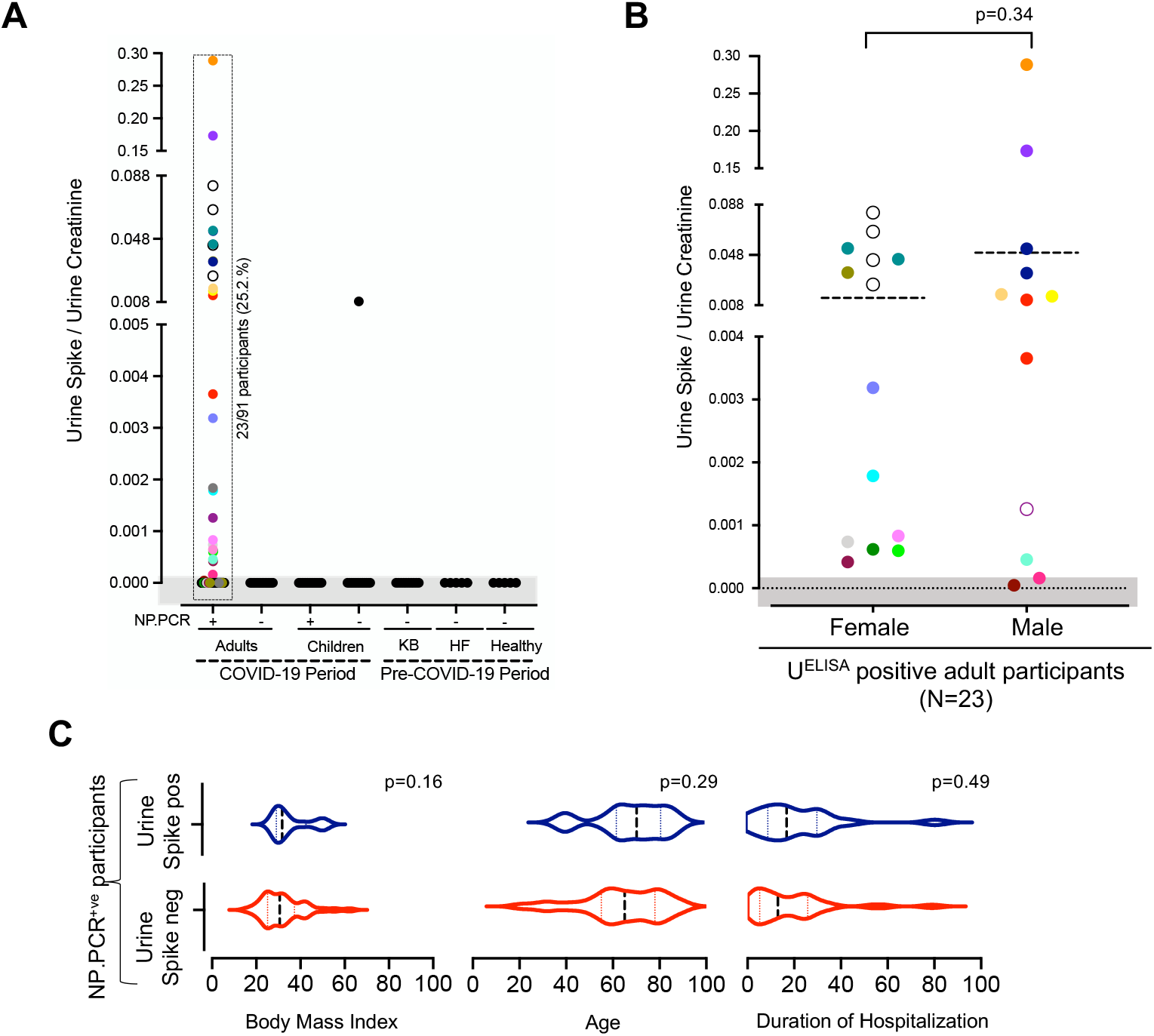
Detection of SARS-CoV-2 in Urine and role of confounding factors. (A) Concentration of SARS-CoV-2 spike protein in urine, measured using Capture ELISA. Multiple urine samples from the same individual are color matched. Multiple urine samples from the same individual are color matched (B-C) Effect of gender, BMI and duration of hospitalization on the presence of SARS-CoV-2 spike protein in urine. Dotted lines indicate mean (B) and median [IQR] (C)

Of the 23 COVID-19 patients with spike protein in their urine, 17 provided at least two urine samples during the course of hospitalization (Figure 3A). SARS-CoV-2 spike protein could be detected in urine from day 1 to day 44 post-hospital admission (Figure 3A). However, no correlation could be found between the concentration of SARS-CoV-2 spike protein in the urine and the day urine sample was collected post hospital admission.

**Figure 3.**
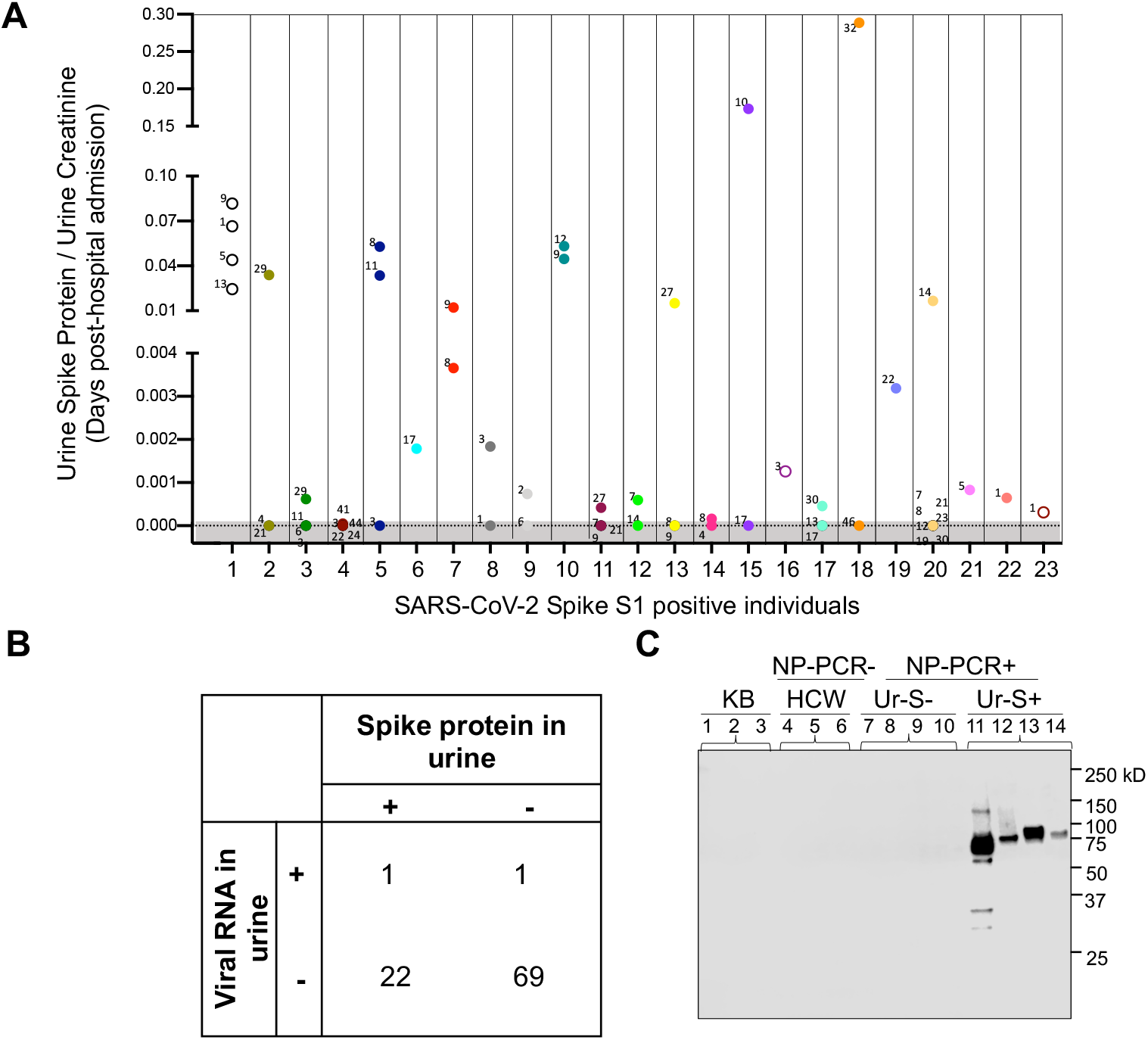
Detection of SARS-CoV-2 in urine samples. (A) Assessment on the concentration of SARS-CoV-2 spike protein in urine using capture ELISA on samples from same individuals collected at different timepoints (shown in days since hospitalization). Multiple urine samples from the same individual are color matched. Data points in the shaded area are below the limit of detection. (B) Detection of viral RNA in urine to determine if the presence of spike protein is due to kidney infection. (C) Detection of SARS-CoV-2 spike protein in urine by Western blot. Lane 1-3 are samples from Kidney Biopsy (KB) participants, Lane 4-6 are from NP-PCR-health care workers, Lane 7-10 are NP-PCR+ individuals who are negative for SARS-CoV-2 spike protein in urine. Lane 11-14 are COVID-19 NP-PCR+ individuals who are positive for SARS-CoV-2 spike protein in urine. A protein fragment size of 78.3 kDa corresponding to SARS-CoV-2 Spike S1 protein is seen in urine spike positive individuals.

To assess whether the presence of spike protein in the urine of a subset of COVID-19 positive individuals is due to the presence SARS-CoV-2 infected cells in this biospecimen, qRT-PCR analysis was conducted on all urine samples from NP-PCR+ individuals using two primer pairs as previously reported.^26^ Out of 93 NP-PCR+ patients, only 2 individuals were positive for viral RNA in urine (∼2%; Figure 3B). Of these 2 positive individuals (1 male and 1 female), only one (female) was positive for both spike protein and viral RNA in the urine.

### Evidence of proteinuria in SARS-CoV-2 infected individuals

To assess whether the presence of the spike protein in the urine of COVID-19 individuals may indicate renal abnormalities caused or exacerbated by viral infection, we analyzed the link between SARS-CoV-2 infection and renal filtration of the human proteins, albumin and cystatin C. Urine samples from 10 patients who underwent kidney biopsy (KB), and collected prior to the pandemic (2015-2018) were included as controls. Our analysis revealed that the concentration of urine albumin among all NP-PCR+ individuals in our cohort was 0.073 [0.019 – 0.276] (median [IQR]; mg/mg of urine creatinine), while urine cystatin C was 0.00014 [0.00008 – 0.00106] (median [IQR]; mg/mg of urine creatinine). Among individuals with SARS-CoV-2 spike protein in the urine, urine albumin concentration was 0.089 [0.016 – 0.299] (median [IQR]; mg/mg of urine creatinine), while urine cystatin C concentration was 0.00012 [0.00007 – 0.00029] (median [IQR]; mg/mg of urine creatinine). The concentration of urine albumin in NP-PCR+ individuals was found to be 0.061 [0.019 – 0.158] (median [IQR]; mg/mg of urine creatinine), whereas that for urine cystatin C was 0.00012 [0.000007 – 0.00050] (median [IQR]; mg/mg of urine creatinine). On the other hand, the concentration of urine albumin in NP-PCR- (HCW) was found to be 0.011 [0.007 – 0.024] (median [IQR]; mg/mg of urine creatinine), whereas that for urine cystatin C was 0.00007 [0.00006 – 0.00008] (median [IQR]; mg/mg of urine creatinine). A total of eight urine samples from six urine ELISA spike positive individuals had albumin concentration greater than 0.3 (mg/mg; Figure 4A, S2A). In total, 22 COVID-19 adult participants (24%; 31 urine samples) had urine albumin greater than 0.3 (mg/mg of urine creatinine; Figure 4A, S2A). Similarly, 18 COVID-19 adult participants (21%; 26 samples) had urine cystatin C concentration greater than 0.0022 [0.001 – 0.003] (median [IQR]; mg/mg of urine creatinine; Figure 4C, S2B). Interestingly, using the cut-off 0.3 of albumin (mg/mg of urine creatinine), which is considered as a marker for AKI,^27^ we found significant association between elevated urine albumin-to-creatinine ratio and concentration of SARS-CoV-2 spike protein in urine (P=0.02; Figure 4B). There were six individuals with elevated urine albumin-to-creatinine ratio greater than 0.3 (mg/mg) who had spike protein in urine, with concentration of 0.0086 [0.00019 – 0.1341]) (median [IQR]; mg/mg of urine creatinine). As shown in Figure 4D, for 2 NP-PCR+ patients (INP.1.019 and INP.1.095) with 3 or 4 urine samples testing positive for SARS-CoV-2 spike protein, our analysis showed no correlation between the levels of urine albumin and cystatin C (normalized to creatinine) and the levels of urine SARS-CoV-2 spike protein (Figure 4D).

**Figure 4.**
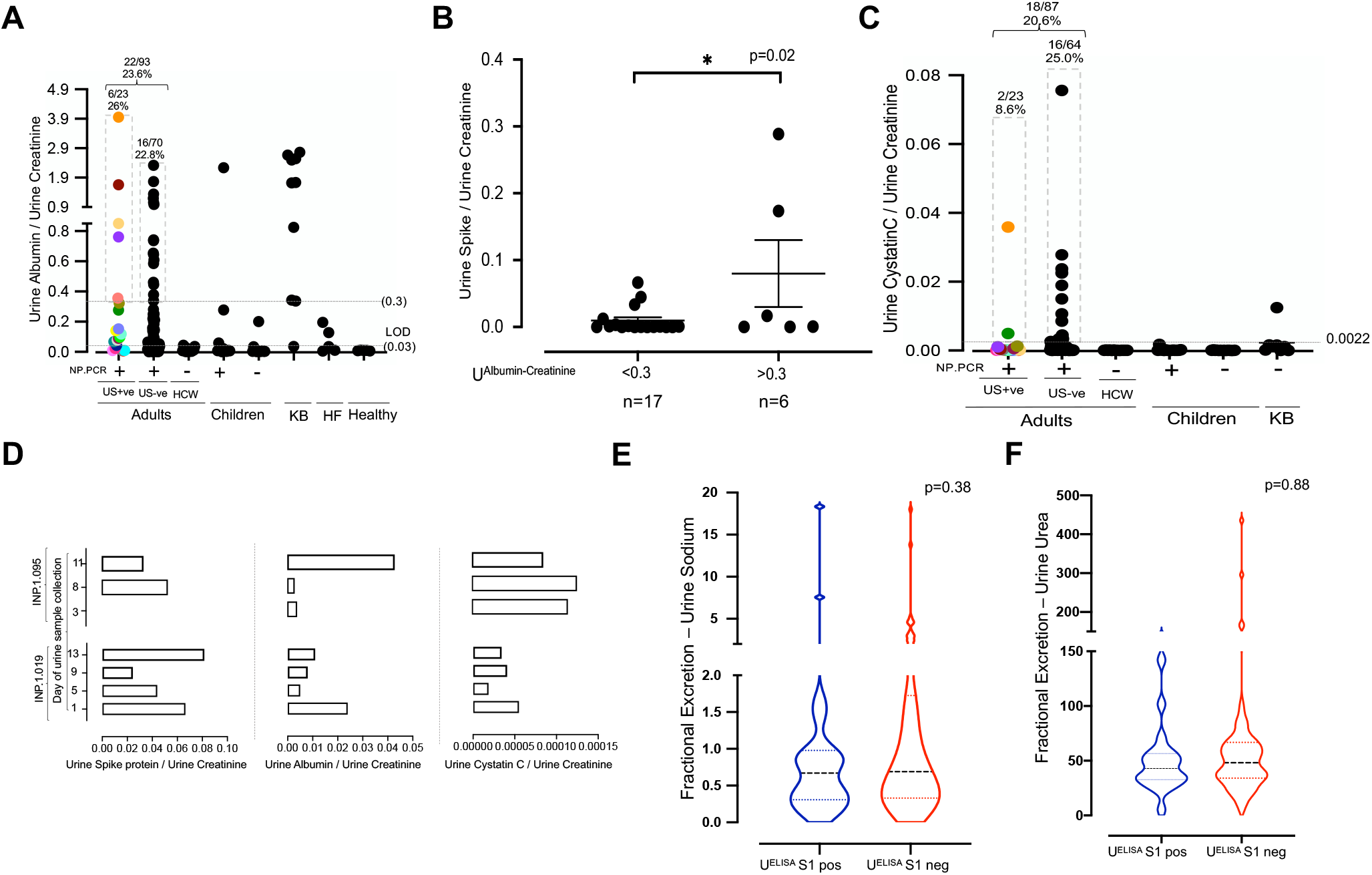
Urine proteomics to determine presence of SARS-CoV-2 spike protein as a biomarker for kidney injury in COVID-19 patients. (A) Concentration of urine albumin (mg/mg of urine creatinine) across one urine sample collected per study participant. Multiple urine samples from the same individual are color matched. (B) Comparison of the concentration of urine spike to albumin concentration (normalized to urine creatinine) greater and less than 0.3. One sample per Ur-S+ participant was considered. The line represents mean ± SEM (C) Concentration of urine cystatin C (mg/mg of urine creatinine) across urine samples, with one collected per study participant. Mean value of cystatin C for KB individuals was determined (0.0022) and used as a cutoff across our study population. Multiple urine samples from the same individual are color matched. (D) Determining correlation between the concentrations of SARS-CoV-2 spike protein, albumin and cystatin C in multiple urine samples from two representative adult NP-PCR+ participants. (E) Fractional excretion of urine sodium. Dashed lines represent median [IQR]. (F) Fractional excretion of urine urea. Dashed lines represent median [IQR].

Previous studies have demonstrated increased level of blood creatinine (>0.3 mg/dL) in individuals with AKI.^28-30^ To determine whether the presence of spike protein in urine was due to AKI, we measured the concentration of creatinine (mg/dL) in sera of individuals that are either positive or negative for spike protein in urine. No significant difference was found between the levels of serum creatinine and the presence or absence of SARS-CoV-2 spike protein in urine among these individuals (P=0.70; Figure S1B). Among individuals with spike protein in urine, the concentration of serum creatinine was 1.319 [0.876 – 1.762] (mean [95%CI]; mg/dL) whereas that of urine spike negative individuals was 1.526 [1.089 – 1.963] (mean [95%CI]; mg/dL). In addition, we analyzed the fractional excretion of sodium (FENa) and urea (FEUrea) levels between samples from urine spike positive and negative individuals, by considering one urine sample per participant (adults N=89; children N=2). No statistically significant difference for both FENa (P=0.38) and FEUrea (P=0.88) between urine spike positive and negative samples (Figure 4E and 4F) could be found. The mean FENa for urine spike positive samples was 1.86 [-0.09 – 3.82] (mean [95%CI]), while that for urine spike negative samples was 1.73 [0.93 – 2.54] (mean [95%CI]). The mean FEUrea for urine spike positive samples was 48.98 [35.06 – 62.90] (mean [95%CI]), while that for urine spike negative samples was 62.13 [45.25 – 79.00] (mean [95%CI]).

Considering the importance of protein size in renal filtration, we used anti-SARS-CoV-2 spike polyclonal antibodies to detect the protein in urine samples by Western blot. As shown in Figure 3C, a band of 78.3 kDa that comigrates with recombinant S1 antigen could be detected in the urine of spike positive SARS-CoV-2 adults. No S1 protein could be detected in the Ur-S-SARS-CoV-2 positive adults, health care workers or individuals whose urine samples were collected prior to the pandemic.

### The presence of spike protein in urine does not correlate with increased levels of viral protein in serum

To assess whether the presence of high levels of SARS-CoV-2 Spike protein in the urine samples from COVID-19 patients may be due to the presence of the protein at high levels in serum, we screened 49 available serum samples from the cohort of 38 COVID-19 patients using spike capture ELISA assay (S^ELISA^) (48 urine samples; N=38). Only 4 samples from 3 individuals showed levels of spike protein in serum above the lower limit of detection (5ng/ml) and none of these were positive in the ELISA assay performed on the urine samples from COVID-19 patients (Figure 5). Together these data suggest that no correlation exists between high levels of spike protein in urine and their serum concentrations.

**Figure 5.**
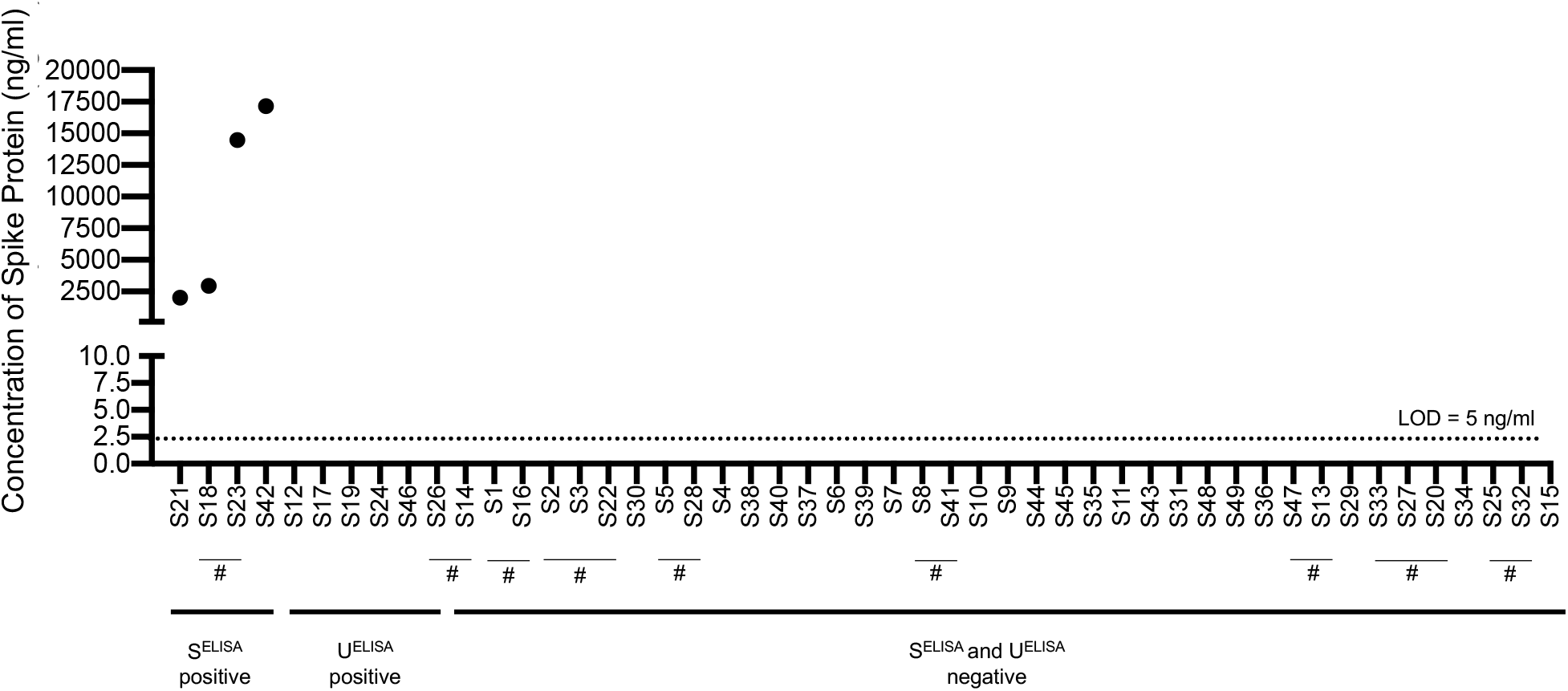
Serum capture ELISA to determine the presence of SARS-CoV-2 spike protein in COVID-19 patients. ELISA assay performed on 49 serum samples collected from 38 COVID-19 patients. # indicates multiple serum samples from the same individual. Dashed line represents the limit of detection (LOD), that is 5ng/ml of spike protein. S^ELISA^ +ve indicates the serum samples positive for spike protein in S^ELISA^. U^ELISA^ +ve indicates the serum samples that are positive for spike protein in U^ELISA^, but are negative in S^ELISA^. S^ELISA^ and in U^ELISA^ -ve samples indicate the serum samples that are negative for spike protein in both S^ELISA^ and U^ELISA^.

## DISCUSSION

In this study, we developed an antigen capture ELISA assay to detect SARS-CoV-2 spike protein and used it to analyze urine samples from a cohort of 152 individuals, including SARS-CoV-2 NP-PCR+ adults and children. While antigen-based detection assays have been reported,^31, 32^ to the best of our knowledge this is the first report of the use of an antigen capture assay to detect spike protein in the urine of SARS-CoV-2 patients and asymptomatic individuals.

Analysis of our urine collection revealed that ∼25% of NP-PCR COVID-19 positive patients shed SARS-CoV-2 spike protein in the urine. In addition, one NP-PCR-child’s urine was found to be positive for SARS-CoV-2 spike protein. In our study, the overall ratio of SARS-CoV-2 spike protein to urine creatinine in adults was 0.033 [0.0096 – 0.0564] (mean [95% CI]; mg/mg of urine creatinine), whereas that of the positive child was 0.0083 (mg/mg of urine creatinine). None of the other urine samples used in our study showed the presence of SARS-CoV-2 spike protein. There was also no correlation between the presence of SARS-CoV-2 spike protein and confounding factors BMI, age, gender and duration of hospitalization. We further assessed whether the presence of spike protein in urine was due to kidney infection, injury or dysfunction in COVID-19 patients. No correlation was observed between the presence of SARS-CoV-2 spike protein in urine and markers of kidney dysfunction including serum creatinine, FENa, FEUrea or cystatin C. However, we noted that the level of SARS-CoV-2 spike protein in urine was higher in patients with albuminuria (Figure 4B). A 2003 study by Chu *et al*. suggested that the development of AKI in SARS-CoV patients was likely to be due to multi-organ failure rather than kidney tropism of the virus.^33^

Interestingly, our analysis of the urine samples by Western blot showed the presence of the expected size (78.3kDa) of the S1 fragment of the spike protein (Figure 3C). In addition, we also observed additional fragments, suggesting proteolysis (Figure 3C). Considering that both spike protein and albumin have molecular weights >60 kDa, it is likely that their release may be the result of similar filtration abnormalities.^34^ Altogether, our data suggest that the presence of spike protein in urine samples of some COVID-19 patients may still be indicative of an unknown or unpredicted kidney injury, most likely spilling of spike protein from serum. Noteworthy, the presence of proteinuria and microscopic hematuria has been associated with greater clinical severity of COVID-19 disease.^20^ The predominant form of kidney injury in COVID-19 seems to be acute tubular injury that might be secondary to cytokine storm or shock. Direct viral infection while present, may only occur in the most severe cases as noted in autopsy studies.^35-37^ Another important finding of this study is the lack of viral RNA in the urine of most NP-PCR+ individuals. Out of 93 NP-PCR+ patients, only 2 individuals were positive for viral RNA in urine (∼2%; Figure 3B). Of these 2 positive individuals (1 male and 1 female), only one (female) was positive for both spike protein and viral RNA in the urine. This suggests that the SARS-CoV-2 spike protein detected in the urine is a direct result of a filtration abnormality rather than a viral infection of the kidney. Whether the presence of viral RNA in the urine of a small percentage of COVID-19 patients is a result of viral shedding or simply due to contamination during urine collection remains to be further evaluated. However, our analysis of urine samples collected at different times from the same individuals during hospitalization do not show a pattern of viral RNA detection that would be consistent with viral RNA shedding in the urine. More importantly, it cannot be ruled out that the urine samples in general have not gone through rigorous accuracy verification or matrix equivalency studies for RT-qPCR as NP or nasal swab samples.

A significant finding in this study is that one child was found to have SARS-CoV-2 spike protein in the urine despite being negative by NP-PCR. Noteworthy, this urine sample was collected on the same day the NP swab was performed for qRT-PCR analysis. One possible explanation is that the child was previously infected and was tested negative due to viral clearance but continues to shed viral spike protein in the urine. Another possibility is that the NP-PCR in this case was a false negative. The ability of the antigen capture assay to detect spike protein in a NP-PCR-individual highlights the need for the development of assays that are not intrusive, are rapid and can be deployed for large scale detection of active infection in the general population and at different times, to prevent continued propagation of the virus. While urine ELISA-based tests are particularly suitable for large scale, repeat and rapid diagnostic campaigns, the fact that only 25% of infected individuals in our study were found to have spike protein in their urine suggests that the sensitivity of the current urine spike-capture assay is not sufficient for population-based screening. Efforts to evaluate the usefulness of this antigen-based assay to detect SARS-CoV-2 infection in other biospecimens such as saliva are warranted.

In conclusion, our data demonstrate that 25% of SARS-CoV-2 infected individuals shed spike protein in the urine. qRT-PCR on urine samples demonstrated that this shedding is neither due to the presence of infected cells in this specimen nor to high levels of this viral protein in the serum. However, it does not preclude the possibility of kidney infection by the virus. Nevertheless, our data highlight possible kidney abnormalities resulting from SARS-CoV-2 infection, the long-term consequences of which require further investigation.

### Ethics statement

This study was approved by Yale Human Research Protection Program Institutional Review Boards (FWA00002571, protocol ID 2000027690). Urine collection from COVID-19 positive and negative children was conducted under protocol IRB20-017503 approved by the Institutional Review Board at Children’s Hospital of Philadelphia (FWA00000459). Informed consent was obtained from all enrolled participants. The pre-COVID-19 urine samples, from Kidney Biopsy participants were provided by Yale BioBank and the study was approved by Yale Human Investigation Committee (number 11110009286). The pre-COVID-19 urine samples from Heart Failure Cohort and healthy individuals were provided by JMT (HIC#1311013065).

## Data Availability

Data available upon request.

## ACKNOWLEDGMENTS

CBM research is supported by grants from National Institute of Health (NIH) and the Steven and Alexandra Cohen Foundation. ARJ is an Investigator in the Pathogenesis of Infectious Diseases of the Burroughs Wellcome Fund, whose work is supported by NIH/NIAID R01AI103280, R21AI144472, and R21AI154370. JMT research was supported by NIH awards (K23HL114868, L30HL115790, R01HL139629, R21HL143092, R01HL128973, R01HL148354). IMPACT received support from the Yale COVID-19 Research Resource Fund. The authors declare no competing financial interests.

## IMPACT Research Team

Kelly Anastasio, Santos Bermejo, Sean Bickerton, Kristina Brower, Edward Courchaine, Rebecca Earnest, John Fournier, Bertie Geng, Laura Glick, Akiko Iwasaki, Chaney Kalinich, Daniel Kim, Maxine Kuang, Eriko Kudo, Sarah Lapidus, Joseph Lim, Alice Lu-Culligan, Irene Matos, Maksym Minasyan, Maura Nakahata, Allison Nelson, Angela Nunez, Camila Odio, Saad Omer, Annsea Park, Mary Petrone. Sarah Prophet, Harold Rahming, Denise Shepard, Erin Silva, Yvette Strong, Pavithra Vijayakumar, Elizabeth B. White, Yexin Yang

## Abbreviations

NP: Nasopharyngeal
ELISA: Enzyme Linked Immunosorbent Assay
HCoV: Human Coronavirus
KB: Kidney Biopsy
AKI: Acute Kidney Infection
U^ELISA^: Urine Capture ELISA
FENa: Fractional Excretion of Sodium
FEUrea: Fractional excretion of Urea
Ur-S+: Urine Spike positive
Ur-S-: Urine Spike negative

## Supplemental Figures

**Figure S1.**
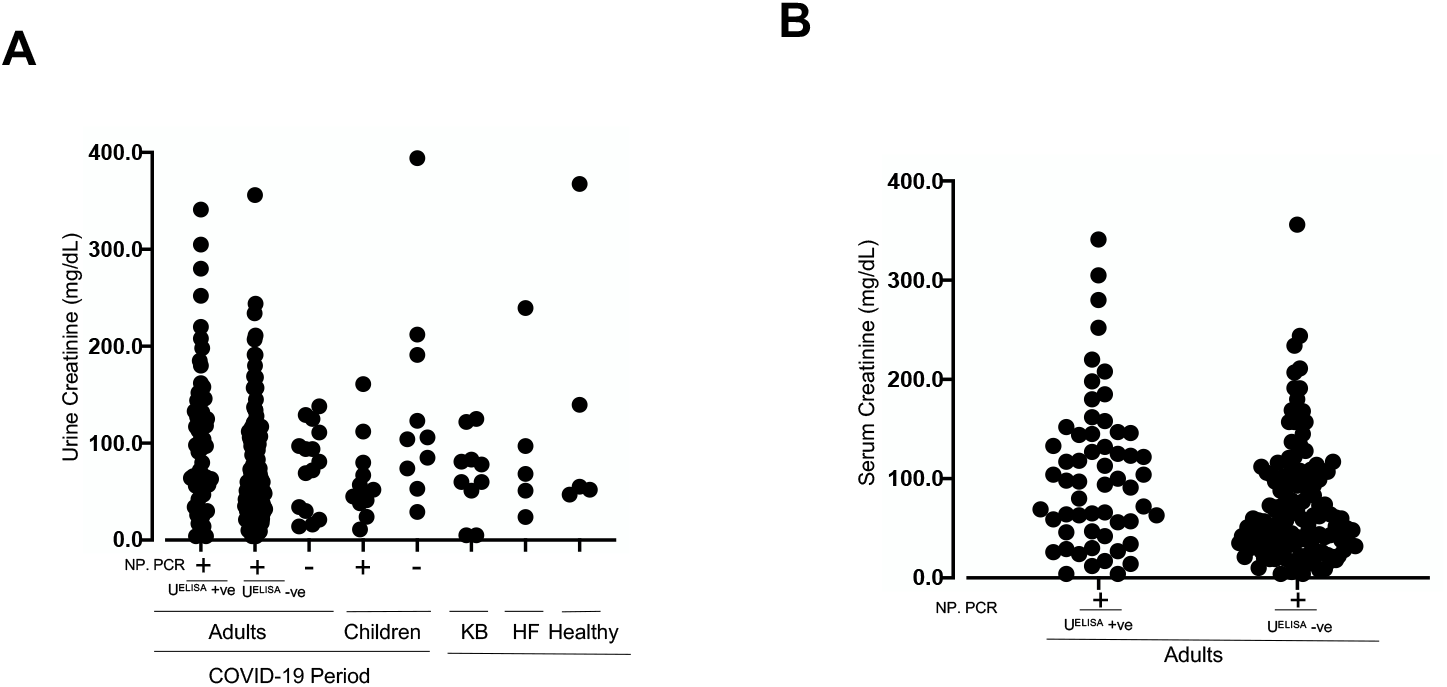
Concentration of urine and serum creatinine across the study population. (A) Comparison of the concentration of urine creatinine across adults (NP-PCR COVID-19 positive and negative participants), children (NP-PCR COVID-19 positive and negative participants) as well as the PreCOVID-19 samples (KB, Heart Failure and Healthy). (B) Comparison of serum creatinine across U^ELISA^ positive and negative participants.

**Figure S2.**
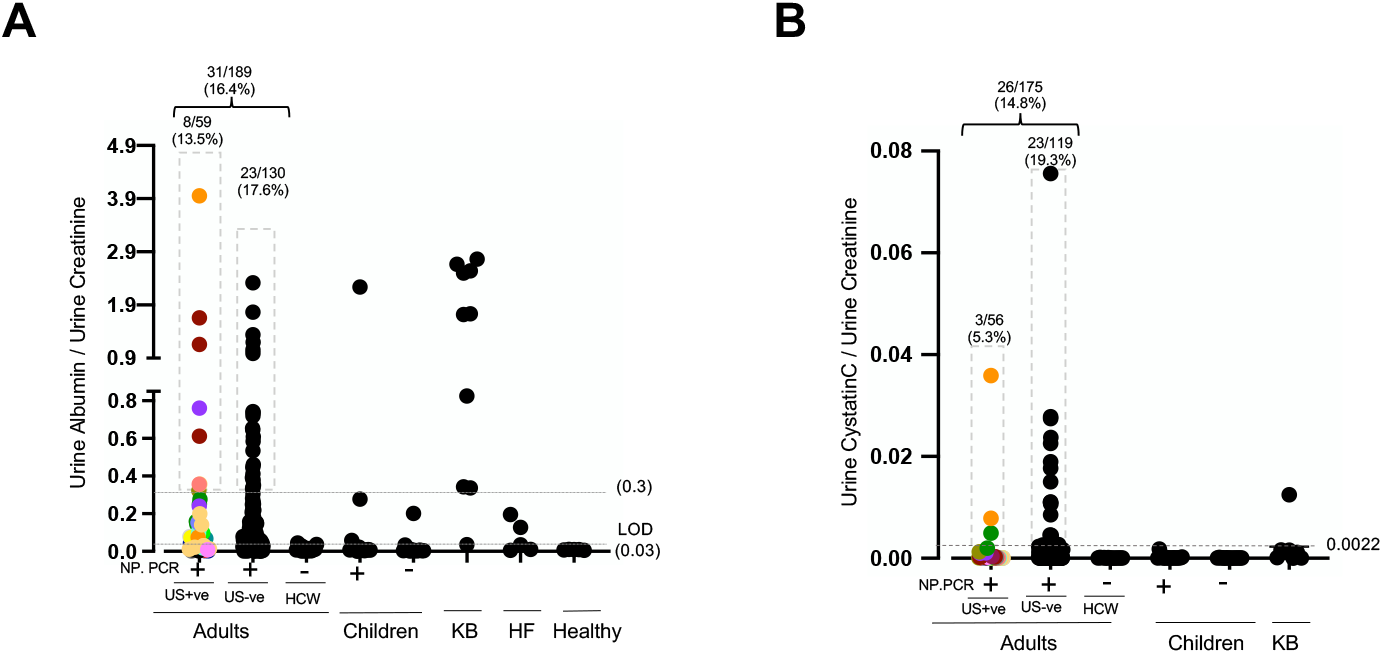
Urine proteomics to determine presence of SARS-CoV-2 spike protein as a biomarker for kidney injury in COVID-19 patients across all the urine samples. Multiple urine samples from the same individual are color matched. (A) Concentration of urine albumin (mg/mg of urine creatinine) across all the urine samples. A value >0.3 (mg/mg of creatinine) was considered as cut-off. (B) Concentration of urine cystatin C (mg/mg of urine creatinine) across all the urine samples. Mean value of cystatin C for KB individuals was determined (0.0022) and used as a cutoff across our study population.

**Figure S3.**
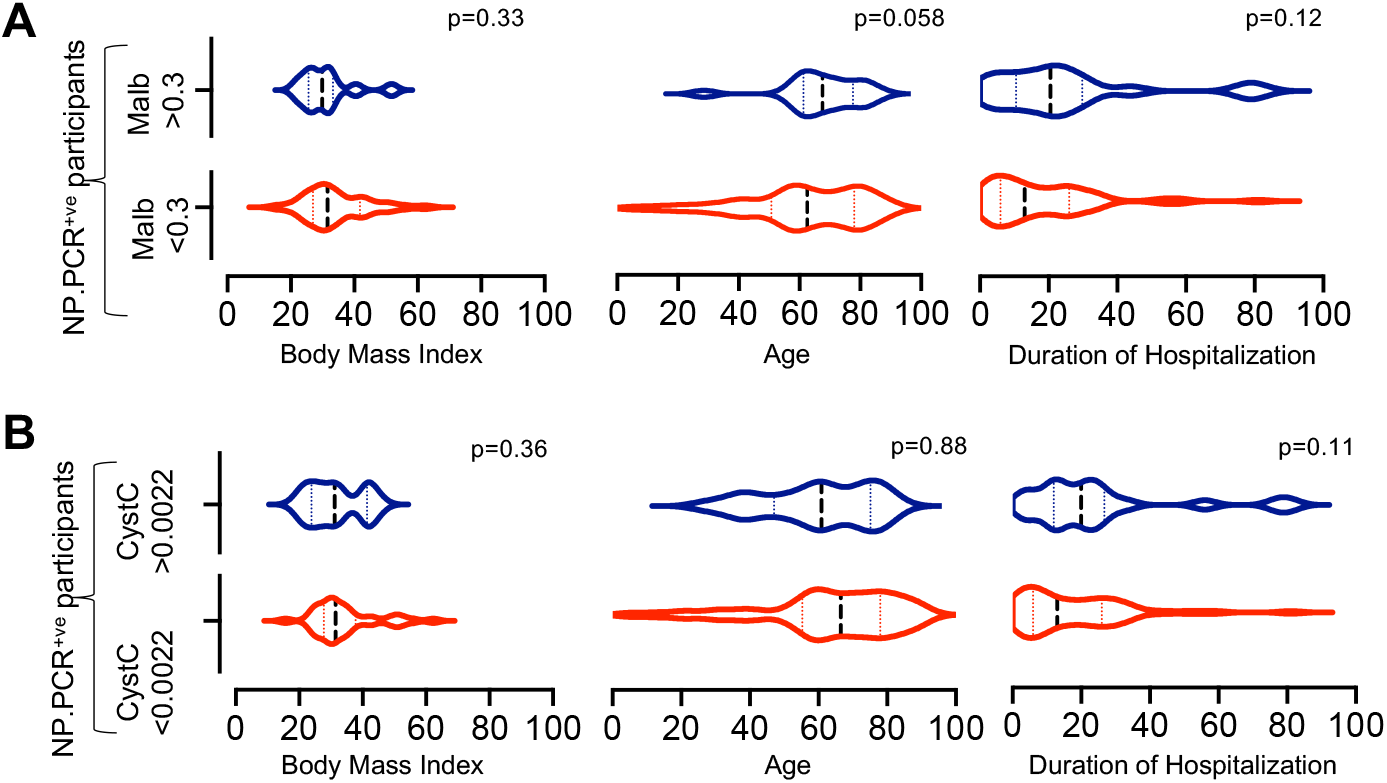
(A,B) Comparison on the role of confounding factors in the elevated levels of albumin in urine. (C,D) Comparison of role of confounding factors in the elevated levels of cystatin C in urine.

**Figure S4.**
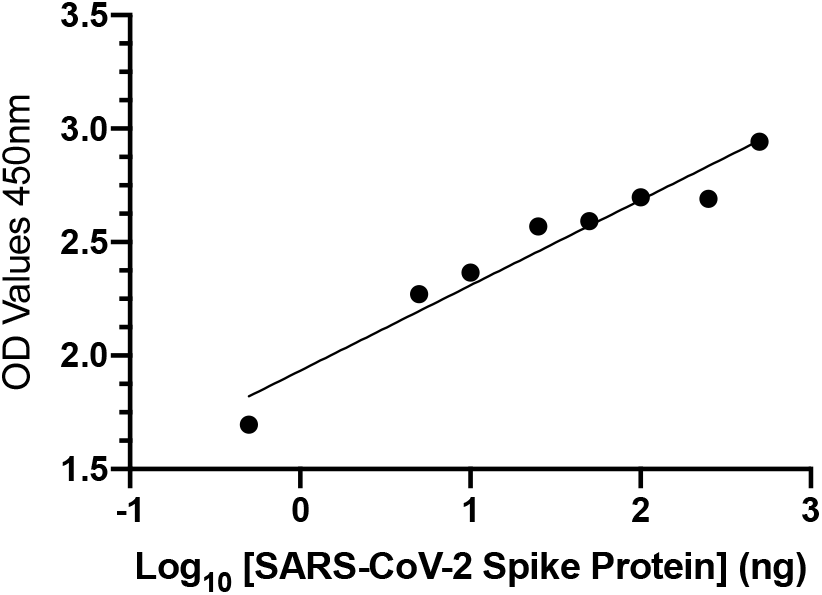
Representative standard curve generated using 5µg/mL SARS-CoV-2 polyclonal anti-spike antibodies.

## Notes

### Competing Interest Statement

The authors have declared no competing interest.

### Author Declarations

This study was approved by Yale Human Research Protection Program Institutional Review Boards (FWA00002571 protocol ID 2000027690). Urine collection from COVID-19 positive and negative children was conducted under protocol IRB20-017503 approved by the Institutional Review Board at Childrens Hospital of Philadelphia (FWA00000459). Informed consent was obtained from all enrolled participants. The pre-COVID-19 urine samples, from Kidney Biopsy participants were provided by Yale BioBank and the study was approved by Yale Human Investigation Committee (number 11110009286). The pre-COVID-19 urine samples from Heart Failure Cohort and healthy individuals were provided by JMT (HIC 1311013065).

